# Health and social inequalities in childhood associate with left ventricular diastolic dysfunction: A life-course EWAS of the 1946 Birth Cohort

**DOI:** 10.1101/2025.09.18.25336102

**Authors:** Matthew Stanley, Eamon Dhall, Constantin-Cristian Topriceanu, Catherine Banks, James C Moon, Marcus Richards, Alun D Hughes, Gabriella Captur

## Abstract

**Background and purpose:** Left ventricular diastolic dysfunction (LVDD) is a key pathophysiological mechanism in heart failure with preserved ejection fraction (HFpEF) but its environmental determinants are poorly understood. We undertook a life-course environment-wide association study (EWAS) of LVDD to test a variety of environmental and biomedical exposures in a high-throughput manner.

**Methods:** Participants were from the Medical Research Council (MRC) National Survey of Health and Development (NSHD, the 1946 British Birth cohort) who had echocardiographic data recorded at 60-64 years. LVDD (outcome) was defined as per the American Society of Echocardiography LVDD guidelines. 255 life-course factors (exposures) were investigated for their association with LVDD. Significant factors were identified using a logistic regression model adjusting for sex, body mass index and socioeconomic position (SEP, dichotomised as manual or non-manual) at a false discovery rate of 5%. Interactions between individual exposures were appraised using exposome correlation globes and matrices.

**Results:** 1616 Participants were included (50.4% men, 21.4% with LVDD) and 21 factors were associated with LVDD (*p*≤0.05) after adjustment. Better childhood cognition (odds ratio 0.86 [95% confidence interval 0.75-0.99]), home conditions (0.90 [0.83-0.97]) and paternal SEP (0.75 [0.56-0.99]) were associated with less LVDD, while childhood home crowding (1.24 [1.09-1.41]), adult consumption of processed pork (1.28 [1.05-1.58]) and sugar (1.24 [1.02-1.50]), and adult systolic hypertension (1.22 [1.06-1.39]) were associated with more LVDD.

**Conclusion:** We have unmasked and rediscovered several exposures throughout the life-course that either protect against or promote, the emergence of LVDD in later life, in particular cognition, housing conditions and diet. Addressing health and social inequalities in childhood might therefore potentially help combat the emerging epidemic of HFpEF.

## INTRODUCTION

Left ventricular diastolic dysfunction (LVDD) can be defined as an intrinsic inability of the LV to expand and fill fully to normal end-diastolic volumes.^1,2^ This results in a reduced cardiac output due to impaired ventricular distensibility and relaxation, and can progress into heart failure with preserved ejection fraction (HFpEF).^3–5^ Indeed, indices of diastolic function (E/e’, tricuspid regurgitation peak velocity, etc.) feature prominently in criteria for HFpEF diagnosis.^6,7^ Identifying risk factors for HFpEF capable of inspiring novel preventative strategies is of great importance.^8,9^

There has been a recent drive by environmental health researchers to explore exposures through environment-wide association studies (EWAS) analogous to how genomics has been tackled through genome-wide association studies.^10–12^ Particularly emphasised has been the concept of the ‘exposome’, defined as the life-course composite of all the exposures (environmental as well as biomedical) of an individual from birth to death.^10,13^ By examining an individual’s life-course exposure, EWAS can unmask associations between specific exposures and population health,^10^ while the exposome allows the evaluation of correlations between individual exposures occurring together.^13^ Our understanding of the life-course exposures which predispose to LVDD is incomplete, with current knowledge mostly confined to traditional cardiometabolic risk factors.^14–16^ We therefore present an EWAS for LVDD, using life-course data from the Medical Research Council (MRC) National Survey of Health and Development (NSHD): the British 1946 birth cohort. Specifically, associations between various environmental and biomedical exposures from birth to older age and echocardiographically-defined LVDD at age 60-64 years were investigated.

## METHODS

### Study population

The MRC NSHD is a prospective birth cohort initially consisting of a nationally representative and age-homogenous sample of 5,326 participants born during one week in March 1946 across the whole of mainland Great Britain. This sample was socially stratified such that study members were single children born to married parents. All children born to non-manual and agricultural parents, plus a random 1-in-4 sample of manual births, were recruited thus containing 2,547 women and 2,815 men. Participants were subjected to regular follow-ups from 1946 to present and assessments collected data on social, environmental, cognitive, mental health and other clinical exposures.

NSHD participants who were still alive with a known address in Britain between 2006-2010, when aged 60-64 years, were invited to a clinic visit that included resting transthoracic echocardiography at six sites across Great Britain. From the original cohort of 5,326, 778 had died, 570 had emigrated, 564 were lost to follow up, and 594 had withdrawn previously. 2,856 participants were invited for assessment between 2006-2010, of which 2,229 accepted: 1,690 attended a clinic and 539 had a home visit.

### Ethical Approval

Written informed consent was obtained from study participants at each wave of follow-up. Ethical approval for the 2006-2010 wave was obtained from the Central Manchester Research Ethics Committee (07/H1008/168) and the Scotland A research Ethics Committee.^17^

### Echocardiography

Echocardiography was offered between 2006-2010 to the 1,690 participants who attended a clinic. Of these, 1,653 accepted and had echocardiography using Vivid I machines (General Electric, USA) of which 1,616 had analysable images. The echocardiographic protocol comprised parasternal long-axis and short-axis views, aortic views, apical five-, four-, three- and two-chamber views and four-chamber tissue Doppler. A blinded analysis was then performed using EchoPac software (General Electric, USA) by three experienced readers according to American Society of Echocardiography (ASE)/European Association of Cardiovascular Imaging (EACVI) guidelines.^18^

Several biomarkers for LVDD were recorded as part of the NSHD echocardiographic protocol. These were the ratio of early (E) to late (A) transmitral filling velocity (E/A), early myocardial relaxation velocity (e’), average E/e’ ratio and left atrial volume indexed to body surface area (LAVI, BSA). E/A was evaluated using pulsed-wave Doppler at the tips of the mitral valve leaflets; e’ using tissue doppler imaging at the septal and lateral mitral annulus; and LAVI through apical 4-chamber and apical 2-chamber views at ventricular end systole.

### Outcome

LVDD was defined by the presence of ≥2 abnormal echocardiographic parameters out of: LAVI (>34 ml/m^2^), mitral E/e’ (>14), septal e’ (<7 cm/s) or lateral e’ (<10 cm/s), and E/A (≥2 or ≤0.8). These cut-off values were in accordance with the ASE guideline for LVDD diagnosis.^18^

### Environmental factors (exposures)

264 Multidimensional factors from within the NSHD dataset were identified as potential EWAS candidates to provide a comprehensive summary of an individual’s life-course environmental and biomedical exposures. As the participants had varying amounts of exposure data recorded throughout their life, the percentage of missing data for each of the 264 exposures was calculated for participants with echocardiographic data.

Factors were organised into 12 domains to comprehensively capture the available NSHD life-course data: Biomarkers, Cardiac, Cognition, Diet, Exercise, General, Mental Health, Pollution, Renal, Respiratory, Smoking, and Social Determinants. Supplementary table 1 details the environmental and biomedical factors found within each domain, with “General” encompassing a small number of variables not described by other domains. To study the life-course effect of an exposure, each study member’s longitudinal data was split into four age categories: childhood (0-18 years), early adulthood (19-44 years), middle age (45-59 years) and older age (60-64 years).

Where possible, objective as opposed to subjective measures of exposures were used to reduce self-report bias. One such example is childhood cognition at age 15 years, determined using the composite of three highly inter-correlated tests to provide a global ability score: the Heim Group Ability Test,^19^ the Watts-Vernon Reading Test,^20^ and a 47-item mathematics test. A minority of exposures were wholly ascertained from self-reporting. This included diet, where a research nurse provided participants detailed instructions on how to complete a comprehensive 7-day prospective food and drink diary, as previously described.^21^ Mental health at ages 53 and 60-64 years was described by the General Health Questionnaire-28 (GHQ), which consists of four 7-item subscales: somatic symptoms, anxiety/insomnia, social dysfunction, and severe depression.^22^

### Covariables

The sex of participants was recorded as male or female (0/1). Height and weight measurements were taken in light, indoor clothing without shoes at age 60-64 years. Height was measured to the nearest millimetre using a portable stadiometer with the head in the Frankfort plane. Weight measurements to the nearest 0.1kg, were taken to calculate body mass index (BMI). Participants’ socio-economic position (SEP) was evaluated at 60-64 years, according to the UK Office of Population Censuses and Surveys Registrar General’s social class, and dichotomized as manual or non-manual (0/1). Where participants had a missing SEP record at age 60-64, the next available SEP was imputed instead.

### Statistical analysis

Statistical analyses were conducted using R version 3.6.0. The normality of continuous data was assessed visually using histograms and by the Shapiro-Wilk test. Continuous sample variables are expressed as mean ± 1 standard deviation (SD); categorical sample variables, as counts and percent.

Significant results were drawn from the complete case analysis and further assessed for validity using multiple imputation (MI) to minimize the bias associated with data missingness. MI was performed using the mice package^23^ to generate missing exposures and covariates under the assumption of missingness at random (MAR).^24^ The imputation model included the outcome (excluding cases with missing outcome), all the exposures, and all covariates. Predictive mean matching MI was used to generate 50 complete data sets using chained equations.^25^

Prior to imputation, skewed exposure variables were transformed to achieve normality. Further, we applied a z-score transformation (adjusting each observation to the mean and scaling by the standard deviation) to compare odds ratios from the many regressions. Similarly, for categorical variables, we made the definition of the referent (0) consistent, defining them as the ‘‘negative/absent/no’’ result of the test. For **Table 1**, differences between LVDD status for normally distributed continuous data were tested using unpaired Student’s t-test, or by Chi-square test for categorical data.

**Table 1.** Baseline and demographic characteristics of the NSHD cohort across different age categories (0-18, 19-44, 45-59, 60-64) according to LVDD status.

| Age Group | Characteristic | No LVDD<br>(n = 1,270) | LVDD<br>(n = 346) | P-value |
| --- | --- | --- | --- | --- |
| 0-18 years | Sex |  |  | 0.999 |
|  | Female - n (%) | 630 (50%) | 171 (49%) |  |
|  | BMI |  |  | 0.215 |
| | Mean ± SD | 20.0 ± 2.6 | 20.2 ± 2.4 | |
|  | SEP |  |  | <0.001 |
|  | Manual - n (%) | 641 (53%) | 192 (58%) |  |
|  | Non-Manual - n (%) | 559 (47%) | 138 (42%) |  |
| 19-44 years | BMI |  |  | <0.001 |
| | Mean ± SD | 23.5 ± 3.1 | 24.3 ± 3.4 | |
|  | SEP |  |  | <0.001 |
|  | Manual - n (%) | 262 (21%) | 103 (30%) |  |
|  | Non-Manual - n (%) | 981 (79%) | 236 (70%) |  |
| 45-59 years | BMI |  |  | <0.001 |
| | Mean ± SD | 26.7 ± 4.2 | 27.9 ± 4.8 | |
|  | SEP |  |  | <0.001 |
|  | Manual - n (%) | 272 (22%) | 106 (31%) |  |
|  | Non-Manual - n (%) | 988 (78%) | 238 (69%) |  |
| 60-64 years | BMI |  |  | <0.001 |
| | Mean ± SD | 27.3 ± 4.5 | 28.8 ± 4.8 | |
|  | SEP |  |  | 0.035 |
|  | Manual - n (%) | 167 (31%) | 61 (41%) |  |
|  | Non-Manual - n (%) | 367 (69%) | 88 (69%) |  |
|  | LV ejection fraction |  |  | <0.001 |
| | Mean ± SD | 64.7 ± 7.5 | 62.9 ± 8.65 | |
|  | n | 1132 | 306 |  |
|  | LV mass index |  |  | <0.001 |
| | Mean ± SD | 111.8 ± 34.8 | 127.7 ± 49.3 | |
|  | n | 909 | 262 |  |

Abbreviations: BMI = body mass index, CI = 95% confidence interval, LVDD = left ventricular diastolic dysfunction, SD = Standard deviation, SEP = Socioeconomic position.

Multivariable logistic regression analysis, adjusting for sex, BMI, and SEP at age 60-64 or 53 years, was performed to examine associations between the 255 environmental factors and LVDD at 60-64 years. When MI was used, regression coefficient estimates, and their 95% confidence intervals (CI) were calculated for each of the 50 datasets and combined using Rubin’s rule. As the population was age-homogenous and essentially all White British, adjustment for age and ethnicity was not required. “Manhattan-style” plots were generated to visualise significant exposures, with *p*-value ≤0.05 being considered significant.

The false discovery rate (FDR) was calculated using the Benjamini-Hochberg (BH) procedure to control for type I error due to multiple hypotheses testing in associating environmental factors to LVDD. An FDR cut-off of 5% was applied. Adjusted associations which were BH significant were thus retained and became the subject of this study’s conclusions.

All the analysed environmental factors were used to define exposome correlation globes to understand intra-and inter-domain correlations between exposures.^13^ One globe was created for each age category using Pearson’s correlation coefficient to test for exposure-outcome associations. To aid the interpretation of intra-domain correlations between exposures, a corresponding exposome matrix was also generated for each age category.^26^ We conducted sensitivity analyses to test the validity and sensitivity of our final estimates by: i) redoing correlations using Spearman instead of Pearson correlations and showing that similar correlation coefficients were obtained; ii) redoing correlations using the imputed dataset instead of the complete case analysis; iii) checking for reverse directionality, or association of exposure due to heart disease diagnosis. To attempt to account for reverse directionality, we recomputed our models omitting individuals who had been diagnosed with heart disease (myocardial infarction, heart failure, cardiomyopathy, aortic stenosis etc). We then refitted our final models with individuals only showing echocardiographic evidence of LVDD without other known prior heart disease diagnosis.

## RESULTS

### Study population characteristics

1616 individuals were included in the study with a mean age of 63±1.1 years. Of these, 346 were classified as having LVDD at age 60-64 years. The median percentage of missingness per exposure was 11.2% (first quartile 6.5% and third quartile 26.6%). None of the participants had complete data on all exposures, yet for 88.2% of individuals <30% of exposure variables had missing values.

Table 1. compares the clinicodemographic characteristics of study members with and without LVDD. In terms of SEP, participants with LVDD were more likely to have had a manual occupation. Those with LVDD had higher mean BMI across all age groups, except 0-18 years, compared with those without. LV ejection fraction was significantly lower and LV mass index higher, in persons with LVDD compared to those without. 801 (49.6%) of all participants were female, and we observed no significant difference in LVDD prevalence between the two sexes.

### Environment-wide associations with LVDD

Figure 1. shows the Manhattan plot summarising multivariable logistic regression results for the 255 environmental exposures, having adjusted for sex, BMI and SEP at age 60-64. Of these, 21 environmental factors retained significant association with LVDD following FDR adjustment to 5% (**Table 2**).

**Table 2.** Significant environmental factors associated with LVDD following multivariable logistic regression adjusting for sex, BMI and SEP at a false discov ery rate of 5%.

| Domain | Variable | Age (years) | P-value | Odds ratio | 95% CI |
| --- | --- | --- | --- | --- | --- |
| Biomarker | Von Willebrand Factor | 60-64 | 0.023 | 1.26 | 1.04 – 1.54 |
| Cardiac | DBP | 53 | 0.001 | 1.25 | 1.09 – 1.43 |
|  | DBP | 60-64 | < 0.001 | 1.42 | 1.22 – 1.66 |
|  | Hypertension | 53 | 0.047 | 1.38 | 1.00 – 1.90 |
|  | Hypertension | 60-64 | 0.0011 | 1.57 | 1.20 – 2.06 |
|  | MAP | 60-64 | < 0.001 | 1.33 | 1.17 – 1.52 |

|  |  |  |  |  |  |
| --- | --- | --- | --- | --- | --- |
|  | SBP | 43 | 0.023 | 1.16 | 1.02 – 1.33 |
|  | SBP | 53 | 0.004 | 1.22 | 1.06 – 1.39 |
|  | SBP | 60-64 | < 0.001 | 1.40 | 1.21 – 1.63 |
| **Cognition** | Cognitive test score | 15 | 0.034 | 0.86 | 0.75 – 0.99 |
| **Diet** | Fruit | 43 | 0.002 | 0.79 | 0.69 – 0.91 |
|  | Processed pork | 53 | 0.019 | 1.28 | 1.05 – 1.58 |
|  | Sugar | 53 | 0.029 | 1.24 | 1.02 – 1.50 |
| **Renal** | Urine creatinine | 60-64 | 0.048 | 0.85 | 0.73 – 1.00 |
| **Social Determinants** | Dwelling upkeep | 11 | 0.04 | 0.78 | 0.61 – 0.99 |
|  | Father's SEP | 11 | 0.040 | 0.75 | 0.56 – 0.99 |
|  | Home ownership | 2-11 | 0.007 | 0.78 | 0.65 – 0.93 |
|  | Household crowding | 2 | 0.001 | 1.24 | 1.09 – 1.41 |
|  | Household crowding | 8 | 0.007 | 1.19 | 1.05 – 1.36 |
|  | Housing quality index | 4 | 0.005 | 0.90 | 0.83 – 0.97 |
|  | Self-employed/Employed | 60-64 | 0.028 | 0.75 | 0.58 – 0.97 |

*Abbreviations: D/SBP = diastolic/systolic blood pressure, GHQ = General Health Questionnaire, MAP = mean arterial pressure, tPA = tissue plasminogen activator. Other abbreviations as in **Table 1**.*

**Figure 1:**
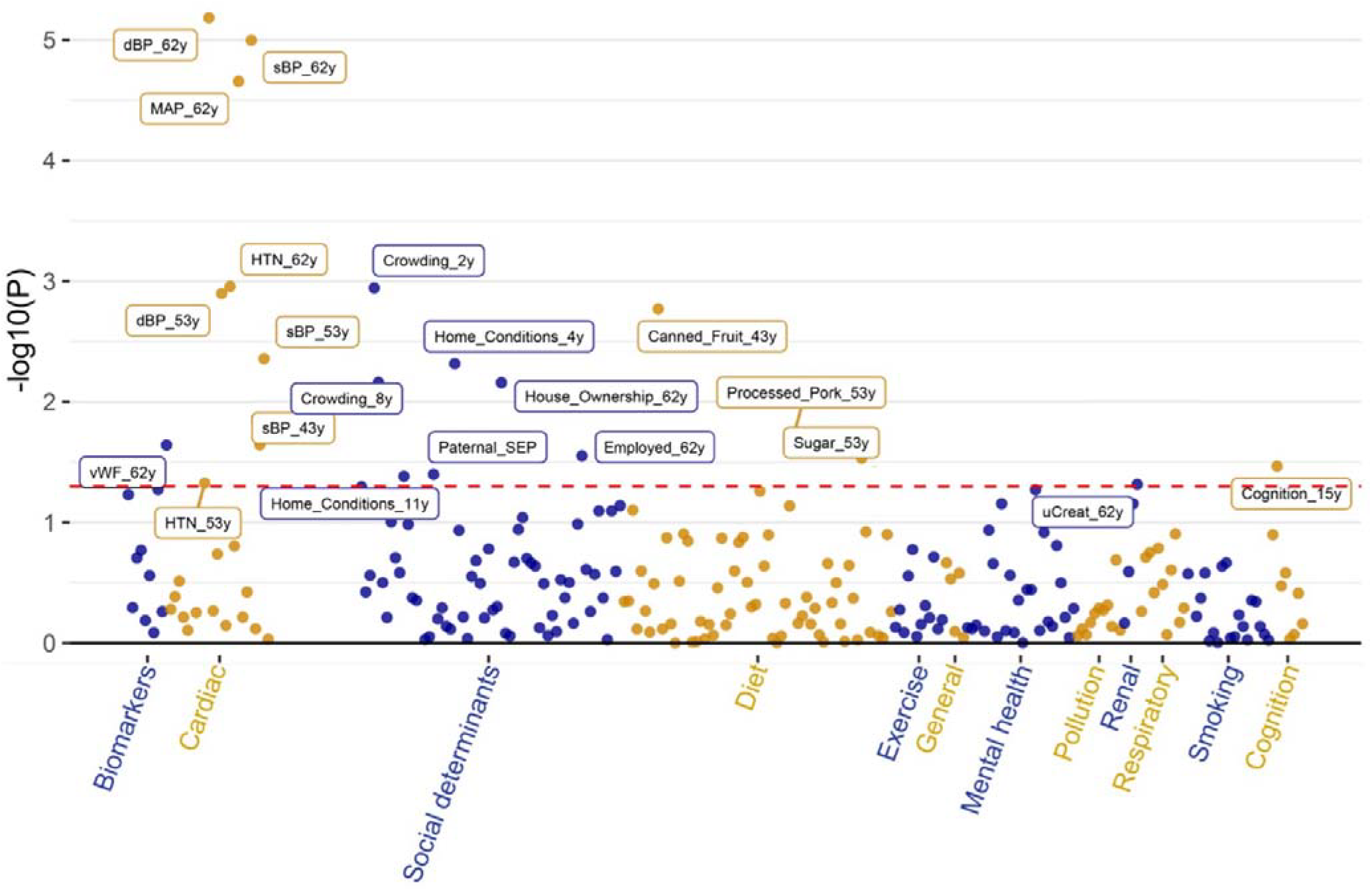
Manhattan plot of multivariable environment-wide associations with left ventricular diastolic dysfunction. Y-axis displays the −log10(*p*-value) of the logistic regression coefficient for each factor tested. X-axis indicates the exposure domains. Factors above the red line are significant (*p*≤0.05). Abbreviations: D/SBP = diastolic/systolic blood pressure, HTN = hypertension, MAP = mean arterial blood pressure, SEP = socioeconomic position, uCreat = urine creatinine, vWF = von Willebrand Factor.

**Figure 2:**
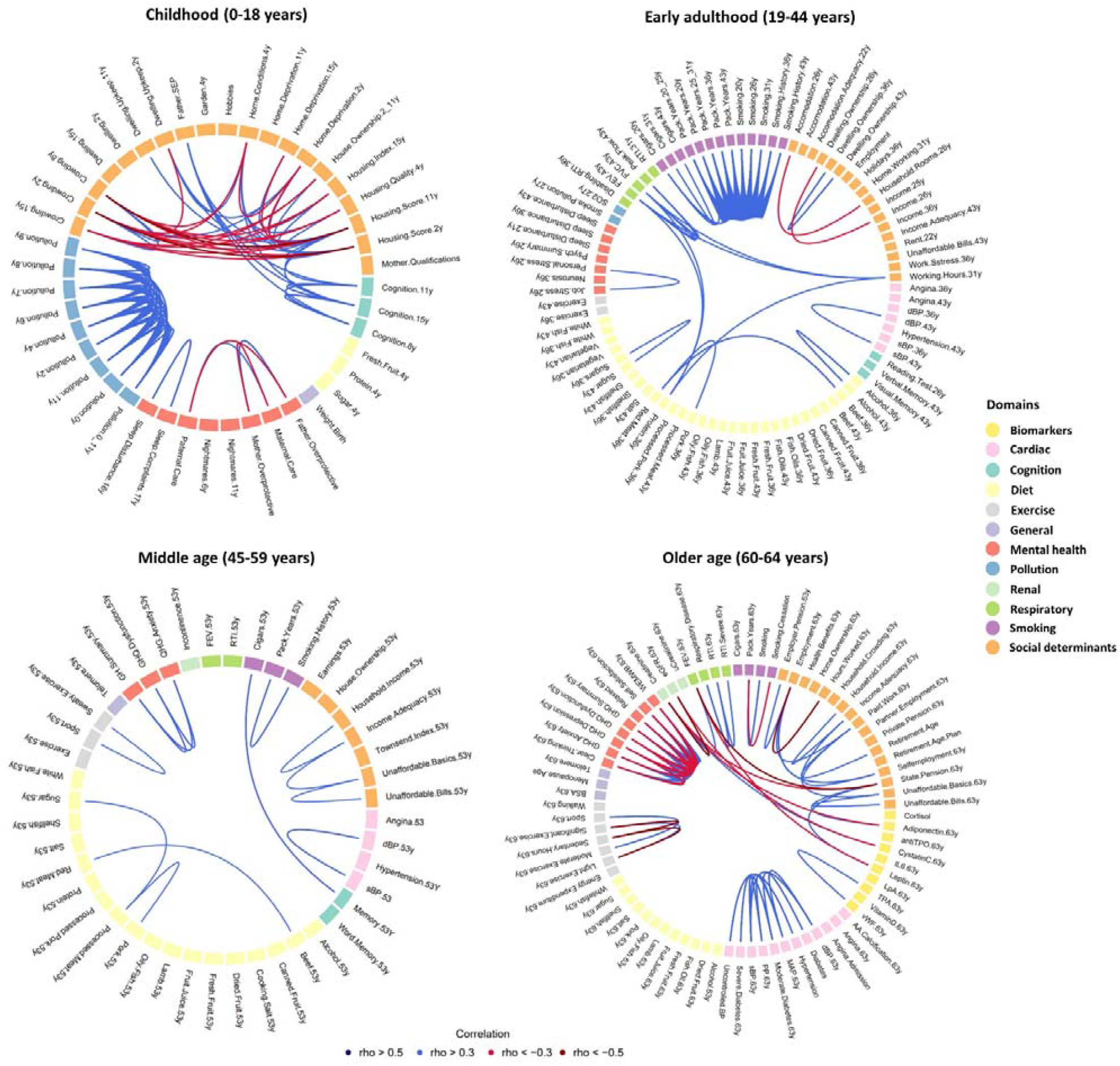
Life course exposome correlation globes for LVDD spanning childhood, early adulthood, middle age and older age. Each line represents a correlation between two exposures. Each domain (key to the right) and correlation is colour coded (key below). Abbreviations: AA.calcification = abdominal aorta calcification, BSA = body surface area, dBP = diastolic blood pressure, FEV = forced expiratory volume, FVC = forced vital capacity, GHQ = general health questionnaire, IL6 = interleukin-6, LpA = lipoprotein A, MAP = mean arterial pressure, PP = pulse pressure, RTI = respiratory tract infection, sBP = systolic blood pressure, SEP = socioeconomic position, TPA = tissue plasminogen activator, TPO = thyroid peroxidase, uCreatinine = urine creatinine, vWF = von Willebrand factor, WEMWB = Warwick-Edinburgh mental wellbeing scale.

### Cardiorenal factors and blood biomarkers

Statistically significant positive associations between systolic and diastolic blood pressure (BP) with LVDD were observed, and these associations were replicated for serial BP measurements between the ages of 43 and 64 years. At 60-64 years, urine creatinine and von Willebrand factor (vWF) levels showed a significant association with LVDD (*p*=0.048; odds ratio [OR] 0.85 [95% CI 0.73-1.00] and *p*=0.023; OR 1.26 [1.04-1.54] respectively).

### Cognitive factors and social determinants

Cognitive ability at 15 years was inversely associated with LVDD in later life (*p*=0.034; OR 0.86 [0.75-0.99]).

Several social determinants relating to housing conditions and SEP during childhood were also significantly associated with LVDD. Of these, the significance of household crowding was replicated at age 2 (*p*=0.001; OR 1.24 [1.09-1.41]) and 8 years (*p*=0.007; OR 1.19 [1.05-1.36]). At 4 years, a holistic range of housing quality markers were indexed, and these revealed an association between poorer housing conditions and LVDD (*p*=0.005; OR 0.90 [0.83-0.97]). Likewise, poorer housing maintenance at 11 years significantly associated with LVDD (*p*=0.040; OR 0.78 [0.61-0.99]). During childhood, father’s non-manual occupation (*p*=0.040; OR 0.75 [0.56-0.99]) and home ownership (*p*=0.007; OR 0.78 [0.65-0.93]), associated with less LVDD. Being employed or self-employed at age 60-64 years was also associated with less LVDD (*p*=0.028; OR 0.75 [0.58-0.97]).

### Dietary factors

Three dietary factors stood out for their association with LVDD. Eating more fruit at age 43 years (*p*=0.002; OR 0.79 [0.69-0.91]) associated with less LVDD, while eating more processed pork (*p*=0.019; OR 1.28 [1.05-1.58]) and more sugar (*p*=0.029; OR 1.24 [1.02-1.50]) at age 53 years, associated with more LVDD.

### The exposome

Many exposures notable for their association with LVDD also displayed correlations with other exposures (**Figure 3** and **Supplementary Figure 1**), with a large proportion of these recorded during childhood. Higher childhood cognition at 15 years was correlated with more advantaged paternal SEP and better composite housing scores at ages 4 and 11 years. Paternal SEP was itself also linked to childhood housing quality at 11 years.

Predictably, many traditional cardiovascular risk factors, including various BP biomarkers, displayed strong intra-domain correlations. This trend was consistent across all age categories.

### Sensitivity analysis

We obtained similar correlations using Spearman instead of Pearson correlations (data not shown) and for the imputed dataset compared to the complete case analysis (**Supplementary Table 3**). Of the 21 significant associations with LVDD in the complete case analysis, 15 remained significant following MI. Next, we considered the possibility of reverse directionality or differential exposure status, due to prior heart disease diagnosis by removing all individuals with known heart disease prior to age 60 years. We then recomputed the effect of exposure, adjusting for sex, BMI and SEP using the remaining individuals who showed echocardiographic evidence of LVDD, but without a known diagnosis of heart disease (**Supplementary Table 4**). Of the exposures that were significant in the main analysis, 19 remained statistically significant after removing these cases. Overall, 13 variables were significant in the complete case analysis and both sensitivity analyses, including childhood cognition and multiple markers of childhood housing quality.

## DISCUSSION

Life-course data from the 1946 British Birth Cohort reveal that besides traditional cardiovascular risk factors, childhood cognitive ability and housing conditions, and adult diet are associated with LVDD in later life. Findings suggest that complex health and social inequalities operating from early life, may be partly responsible for the emerging epidemic of HFpEF.

The exposome is a rapidly developing paradigm and here we executed the first ‘life-course exposome’ for LVDD using longitudinal birth cohort data spanning 7 decades in >1600 British persons. We also sought to uncover the associations between individual exposures, analogous to the “linkage disequilibrium” in genome-wide association studies, by generating exposome correlation globes and matrices. This allowed us to identify “mixtures” or combinations of factors with common routes of exposure, not otherwise possible.

The association between midlife blood pressure and LVDD in later life is well-established and has been previously reported in NSHD, with E/e’ used as the sole surrogate for LVDD.^15^ In the current work, we similarly observed modest increases in the odds of LVDD arising from systolic and diastolic hypertension. Similarly, there are numerous studies describing the association between renal disease and diastolic dysfunction.^27–29^ Our observation that reduced spot urine creatinine levels associate with LVDD should be interpreted cautiously since it was not a 24-hour recording, and estimated glomerular filtration rate did not reach statistical significance.

There is evidence that childhood cognitive ability is inversely correlated with cardiovascular disease^37,38^ and that more time spent in education is inversely associated with incident LVDD.^39^ However, to the best of our knowledge, no study yet has looked at childhood cognition in regard to diastolic function in later life. Our results suggest that higher childhood cognitive ability (at 15 years) is independently protective of LVDD, even after adjustment for adulthood SEP. The exposome globe offers some potential mechanistic insights, given that lower childhood cognition appears to correlate with a lower childhood SEP and poorer composite housing scores. As childhood SEP and housing quality at 4 years were both also independently associated with LVDD, a common underlying mechanism becomes plausible. All three are likely selecting into cardiovascular disease risk, but cognition may additionally be a biomarker for common physiological cause. Occupational SEP across the life-course, including childhood, has been previously associated with echocardiographic markers of LVDD in NSHD.^40^

vWF is a plasma glycoprotein crucial in haemostasis and thrombosis via platelet aggregation and increasing circulatory factor VIII stability.^30^ Its release is stimulated by a variety of triggers often linked with endothelial injury and dysfunction.^31^ Although vWF has been implicated in vascular inflammation, the link with cardiovascular disease is more tenuous.^32–35^ More recently, higher vWF levels alongside other inflammatory biomarkers have been associated with HFpEF, with endothelial dysfunction and inflammation touted as a potential mechanism.^33,36^ Our population-based results suggest the link could extend to LVDD, and should prompt further work to fully elucidate the potential association.

We explored several dietary factors for their association with LVDD. Two of these–consumption of sugar and processed pork in middle age–appear to increase the risk of LVDD, while fruit consumption in early adulthood may be protective. However, a large proportion of dietary variables may have been confounded. The 7-day dietary diaries used in NSHD cannot overcome the problem of self-reporting bias leading to over/underexaggerated reporting of certain foods;^41^ however, the quality and depth of collected data still surpasses many other single timepoint questionnaires.

Our results add to a previous environment-wide association study for LVDD, which identified that age, sex and diabetes were independent predictors.^42^ Rather than a cross-sectional approach used previously, we present a life-course study from birth to older age, and further delineate the influence of environmental factors on LVDD.

Study limitations include the fact that causal relationships cannot be inferred from the discovered exposures. It has been previously shown that NSHD study members who did not have echocardiographic data recorded at age 60-64 years, had higher BMI and resting heart rate, and lower childhood SEP^15,40^ compared to the analytic sample suggesting that our sample was therefore slightly healthier and this could limit the generalisability of findings. Some variables previously associated with LVDD, such as exercise and smoking, failed to show significant association with LVDD in our study. LVDD could have been better defined using the complete ASE diagnostic algorithm, but not all the necessary echocardiographic parameters required for this were available in the NSHD echocardiographic protocol, hence our use of the composite endpoint instead. One biomarker in this endpoint–the E/A ratio–has a known U-shaped relationship with LVDD, meaning that pseudo-normalisation may have attenuated some of our associations, as such persons with modest LVDD may have been missed.

## CONCLUSION

We undertook a prospective life-course EWAS for LVDD. Using exposome correlation globes and matrices, we scrutinised >250 factors across 7 decades of the human life-course and discovered complex mixtures of exposures relevant to LVDD in older age. Amongst these were an intriguing set of novel exposures associating with more LVDD, such as poorer cognition in childhood and diet in adulthood. Findings emphasise the importance of addressing health and social inequalities early in the human life-course, as they might be implicated in exacerbating the global health challenge of HFpEF.

## Supporting information

Supp manuscript

## Data Availability

All data produced in the present study are available upon reasonable request to the authors and available from the LHA office through skylark and Condor portals.

https://nshd.mrc.ac.uk/data-sharing/

