## Supplementary material for "Health and social inequalities in childhood associate with left ventricular diastolic dysfunction: A life-course EWAS of the 1946 Birth Cohort": Supp manuscript

**Table S1:** All 255 exposures analysed in this study, organised into age of exposure and the domain classification.

| **Childhood (0-18 years)** | | **Young adulthood (19-44 years)** | | **Middle age (45-59 years)** | | **Older age (60-64 years)** | |
| --- | --- | --- | --- | --- | --- | --- | --- |
| Domain | Exposure | Domain | Exposure | Domain | Exposure | Domain | Exposure |
| General | Weight at birth | Respiratory | RTI 31y | Respiratory | RTI 53y | Respiratory | RTI Severe 63y |
| Cognition | Cognition 11y | Respiratory | Disabling RTI 36y | Respiratory | FEV 53y | Respiratory | RTI 63y |
| Cognition | Cognition 8y | Respiratory | FEV 43y | Cardiac | Angina 53 | Respiratory | FEV 63y |
| Cognition | Cognition 15y | Respiratory | FVC 43y | Cardiac | dBP 53y | Respiratory | Respiratory Disease 63y |
| Social  Determinants | Father SEP | Respiratory | Peak Flow.43y | Cardiac | sBP 53y | Cardiac | Angina 63y |
| Social  Determinants | Housing Quality.4y | Cardiac | Angina 36y | Cardiac | Hypertension 53y | Cardiac | Angina Admission |
| Social  Determinants | Housing Score.2y | Cardiac | Angina 43y | Cognition | Memory 53y | Cardiac | Moderate Diabetes 63y |
| Social  Determinants | Housing Score.11y | Cardiac | dBP 36y | Cognition | Word Memory 53y | Cardiac | Diabetes diagnosis 63y |
| Social  Determinants | Housing Index.15y | Cardiac | dBP 43y | Social determinants | Townsend Index 53y | Cardiac | Severe Diabetes 63y |
| Social Determinants | Dwelling 2y | Cardiac | sBP 36y | Social determinants | House Ownership 53y | Cardiac | Abdominal Aorta Calcification 63y |
| Social Determinants | Dwelling 15y | Cardiac | sBP 43y | Social determinants | Earnings 53y | Cardiac | dBP 63y |
| Social Determinants | House Ownership 2-11y | Cardiac | Hypertension 43y | Social determinants | Household Income 53y | Cardiac | sBP 63y |
| Social Determinants | Hobbies | Cognition | Verbal Memory 43y | Social determinants | Unaffordable Bills 53y | Cardiac | PP 63y |
| Social Determinants | Home Conditions 4y | Cognition | Visual Memory 43y | Social determinants | Income Adequacy 53y | Cardiac | MAP 63y |
| Social Determinants | Dwelling Upkeep 2y | Cognition | Reading Test 26y | Social determinants | Unaffordable Basics 53y | Cardiac | Hypertension 63y |
| Social Determinants | Dwelling Upkeep 11y | Social determinants | Working Hours 31y | Mental health | GHQ Summary 53y | Cardiac | Uncontrolled BP |
| Social Determinants | Home Deprivation 2y | Social determinants | Home Working 31y | Mental health | GHQ Anxiety 53y | General | BSA 63y |
| Social Determinants | Home Deprivation 11y | Social determinants | Accommodation 26y | Mental health | GHQ Dysfunction 53y | General | Menopause Age |
| Social Determinants | Home Deprivation 15y | Social determinants | Accommodation 43y | Renal | Incontinence 53y | General | Telomere 63y |
| Social Determinants | Garden 4y | Social determinants | Dwelling Ownership 36y | Exercise | Exercise53y | Social determinants | Partner Employment 63y |
| Social Determinants | Crowding 2y | Social determinants | Dwelling Ownership 43y | Exercise | Sweaty Exercise 53y | Social determinants | Hours Worked 63y |
| Social Determinants | Crowding 8y | Social determinants | Dwelling Ownership 26y | Exercise | Sport 53y | Social determinants | Retirement Age Plan |
| Social Determinants | Crowding 15y | Social determinants | Work Stress 36y | Diet | Alcohol 53y | Social determinants | Retirement Age |
| Social Determinants | Mother Qualifications | Social determinants | Holidays 36y | Diet | Salt 53y | Social determinants | Employment 63y |
| Mental health | Sleep Disturbance.16y | Social determinants | Income 36y | Diet | Cooking Salt 53y | Social determinants | Paid Work 63y |
| Mental health | Nightmares 6y | Social determinants | Income 25y | Diet | Protein 53y | Social determinants | Household Crowding 63y |
| Mental health | Nightmares 11y | Social determinants | Unaffordable Bills 43y | Diet | Oily Fish 53y | Social determinants | Home Ownership 63y |
| Mental health | Sleep Complaints 17y | Social determinants | Income Adequacy 43y | Diet | Shellfish 53y | Social determinants | Household Income 63y |
| Mental health | Maternal Care | Social determinants | Accommodation Adequacy 22y | Diet | White Fish 53y | Social determinants | Unaffordable Bills 63y |
| Mental health | Paternal Care | Social determinants | Rent 22y | Diet | Fruit Juice 53y | Social determinants | Health Benefits 63y |
| Mental health | Father Overprotective | Social determinants | Household Rooms 26y | Diet | Canned Fruit 53y | Social determinants | State Pension 63y |
| Mental health | Mother Overprotective | Social determinants | Income 26y | Diet | Dried Fruit 53y | Social determinants | Private Pension 63y |
| Diet | Protein 4y | Social determinants | Employment | Diet | Fresh Fruit 53y | Social determinants | Income Adequacy 63y |
| Diet | Fresh Fruit 4y | Mental health | Sleep Disturbance 36y | Diet | Lamb 53y | Social determinants | Self-employment/ employment 63y |
| Diet | Sugar 4y | Mental health | Sleep.Disturbance 43y | Diet | Pork 53y | Social determinants | Employer Pension 63y |
| Pollution | Pollution 11y | Mental health | Sleep.Disturbance 21y | Diet | Beef 53y | Social determinants | Unaffordable Basics 63y |
| Pollution | Pollution 0y | Mental health | Neurosis 36y | Diet | Red Meat 53y | Mental health | WEMWB 63y |
| Pollution | Pollution 2y | Mental health | Personal Stress 26y | Diet | Processed Meat 53y | Mental health | GHQ Summary 63y |
| Pollution | Pollution 4y | Mental health | Psych Summary 26y | Diet | Processed Pork 53y | Mental health | GHQ Depression 63y |
| Pollution | Pollution 6y | Mental health | Job Stress 26y | Diet | Sugar 53y | Mental health | GHQ Anxiety 63y |
| Pollution | Pollution 7y | Exercise | Exercise 43y | General | Telomere 53y | Mental health | GHQ Dysfunction 63y |
| Pollution | Pollution 8y | Exercise | Exercise 36y | Smoking | Smoking History 53y | Mental health | Relaxed 63y |
| Pollution | Pollution 9y | Diet | Alcohol 36y | Smoking | Pack Years 53y | Mental health | Clear Thinking 63y |
| Pollution | Pollution 0-11y | Diet | Alcohol 43y | Smoking | Cigars 53y | Mental health | Self-Satisfaction 63y |
|  |  | Diet | Vegetarian 36y |  |  | Exercise | Sedentary Hours 63y |
|  |  | Diet | Salt 43y |  |  | Exercise | Light Exercise 63y |
|  |  | Diet | Protein 36y |  |  | Exercise | Significant Exercise 63y |
|  |  | Diet | White Fish 36y |  |  | Exercise | Sport 63y |
|  |  | Diet | Shellfish 36y |  |  | Exercise | Energy Expenditure 63y |
|  |  | Diet | Oily Fish 36y |  |  | Exercise | Walking 63y |
|  |  | Diet | Fish Oils 36y |  |  | Exercise | Moderate Exercise 63y |
|  |  | Diet | Oily Fish 43y |  |  | Diet | Alcohol 63y |
|  |  | Diet | Shellfish 43y |  |  | Diet | Salt 63y |
|  |  | Diet | White Fish 43y |  |  | Diet | Whitefish 63y |
|  |  | Diet | Fish Oils 43y |  |  | Diet | Shellfish 63y |
|  |  | Diet | Fruit Juice 36y |  |  | Diet | Oily Fish 63y |
|  |  | Diet | Fresh Fruit 36y |  |  | Diet | Fish Oil 63y |
|  |  | Diet | Dried Fruit 36y |  |  | Diet | Fruit Juice 63y |
|  |  | Diet | Canned Fruit 36y |  |  | Diet | Fresh Fruit 63y |
|  |  | Diet | Canned Fruit 43y |  |  | Diet | Dried Fruit 63y |
|  |  | Diet | Dried Fruit 43y |  |  | Diet | Lamb 63y |
|  |  | Diet | Fresh Fruit 43y |  |  | Diet | Pork 63y |
|  |  | Diet | Fruit Juice 43y |  |  | Diet | Sugar 63y |
|  |  | Diet | Sugars 36y |  |  | Smoking | Smoking |
|  |  | Diet | Beef 36y |  |  | Smoking | Smoking Cessation |
|  |  | Diet | Pork 36y |  |  | Smoking | Pack Years 63y |
|  |  | Diet | Red Meat 36y |  |  | Smoking | Cigars 63y |
|  |  | Diet | Processed Pork 36y |  |  | Biomarkers | antiTPO 63y |
|  |  | Diet | Beef 43y |  |  | Biomarkers | CystatinC 63y |
|  |  | Diet | Lamb 43y |  |  | Biomarkers | VitaminD 63y |
|  |  | Diet | Processed Meat 43y |  |  | Biomarkers | vWF 63y |
|  |  | Diet | Sugar 43y |  |  | Biomarkers | TPA 63y |
|  |  | Pollution | Smoke Pollution 27y |  |  | Biomarkers | LpA 63y |
|  |  | Pollution | Sulphur dioxide 27y |  |  | Biomarkers | Leptin 63y |
|  |  | Smoking | Smoking 20y |  |  | Biomarkers | IL6 63y |
|  |  | Smoking | Smoking 31y |  |  | Biomarkers | Adiponectin 63y |
|  |  | Smoking | Smoking 26y |  |  | Biomarker | Cortisol 63y |
|  |  | Smoking | Smoking History 43y |  |  | Renal | Creatinine 63y |
|  |  | Smoking | Smoking History 36y |  |  | Renal | eGFR 63y |
|  |  | Smoking | Pack Years 20y |  |  | Renal | Urine creatinine 63y |
|  |  | Smoking | Pack Years 20-25y |  |  |  |  |
|  |  | Smoking | Pack Years 25-31y |  |  |  |  |
|  |  | Smoking | Pack Years 43y |  |  |  |  |
|  |  | Smoking | Pack Years 36y |  |  |  |  |
|  |  | Smoking | Cigars 43y |  |  |  |  |
|  |  | Smoking | Cigars 31y |  |  |  |  |

*Abbreviations: BSA = body surface area, dBP = diastolic blood pressure, FEV = forced expiratory volume, FVC = forced vital capacity, GHQ = general health questionnaire, IL6 = interleukin-6, LpA = lipoprotein A, MAP = mean arterial pressure, PP = pulse pressure, RTI = respiratory tract infection, sBP = systolic blood pressure, SEP = socioeconomic position, TPA = tissue plasminogen activator, TPO = thyroid peroxidase, vWF = von Willebrand factor, WEMWB = Warwick-Edinburgh mental wellbeing scale.*

**Table S2**: Table of references detailing the methodology for significant exposures in this study.

| Biomarkers | Murray, E., Hardy, R., Hughes, A., Wills, A., Sattar, N., Deanfield, J., Kuh, D., & Whincup, P. (2015). Overweight across the life course and adipokines, inflammatory and endothelial markers at age 60&ndash;64 years: Evidence from the 1946 birth cohort. *International Journal of Obesity*, *39*, 1010–1018. https://doi.org/10.1038/ijo.2015.19 |
| --- | --- |
| Blood pressure | Ghosh, A. K., Hughes, A. D., Francis, D., Chaturvedi, N., Pellerin, D., Deanfield, J., Kuh, D., Mayet, J., & Hardy, R. (2016). Midlife blood pressure predicts future diastolic dysfunction independently of blood pressure. *Heart*, *102*(17), 1380–1387. https://doi.org/10.1136/heartjnl-2015-308836 |
| Childhood cognition | Richards, M., & Wadsworth, M. E. J. (2004). Long term effects of early adversity on cognitive function. *Archives of Disease in Childhood*, *89*(10), 922–927. https://doi.org/10.1136/ADC.2003.032490 |
| Childhood socioeconomic factors | Muthuri, S. G., Kuh, D., & Cooper, R. (2018). Longitudinal profiles of back pain across adulthood and their relationship with childhood factors: Evidence from the 1946 British birth cohort. *Pain*, *159*(4), 764–774. <https://doi.org/10.1097/J.PAIN.0000000000001143>  Allinson, J. P., Hardy, R., Donaldson, G. C., Shaheen, S. O., Kuh, D., & Wedzicha, J. A. (2017). Combined impact of smoking and early-life exposures on adult lung function trajectories. *American Journal of Respiratory and Critical Care Medicine*, *196*(8), 1021–1030. https://doi.org/10.1164/RCCM.201703-0506OC |
| Diet and smoking | Maddock, J., Ziauddeen, N., Ambrosini, G. L., Wong, A., Hardy, R., & Ray, S. (2018). *Adherence to a Dietary Approaches to Stop Hypertension (DASH)-type diet over the life course and associated vascular function: a study based on the MRC 1946 British birth cohort*. https://doi.org/10.1017/S0007114517003877 |
| Smoking | Murray, E., Hardy, R., Hughes, A., Wills, A., Sattar, N., Deanfield, J., Kuh, D., & Whincup, P. (2015). Overweight across the life course and adipokines, inflammatory and endothelial markers at age 60&ndash;64 years: Evidence from the 1946 birth cohort. *International Journal of Obesity*, *39*, 1010–1018. https://doi.org/10.1038/ijo.2015.19 |
| Urine creatinine | Silverwood, R. J., Pierce, M., Hardy, R., Sattar, N., Whincup, P., Ferro, C., Savage, C., Kuh, D., & Nitsch, D. (2013). Low birth weight, later renal function, and the roles of adulthood blood pressure, diabetes, and obesity in a British birth cohort. *Kidney International*, *84*, 1262–1270. https://doi.org/10.1038/ki.2013.223 |

|  |  |  | **Complete Case Analysis** | | | **Multiple Imputation** | | |
| --- | --- | --- | --- | --- | --- | --- | --- | --- |
| **Domain** | **Variable** | **Age (years)** | **P-value** | **Odds ratio** | **95% CI** | **P-value** | **Odds ratio** | **95% CI** |
| **Biomarker** | Von Willebrand Factor | 60-64 | **0.023** | 1.26 | 1.04 – 1.54 | 0.051 | 1.17 | 1.00-1.39 |
| **Cardiac** | Hypertension | 53 | **0.047** | 1.38 | 1.00 – 1.90 | 0.138 | 1.26 | 0.92-1.71 |
|  | Hypertension | 60-64 | **0.001** | 1.57 | 1.20 – 2.06 | **0.008** | 1.42 | 1.09-1.83 |
|  | DBP | 53 | **0.001** | 1.25 | 1.09 – 1.43 | **0.001** | 1.23 | 1.09-1.40 |
|  | DBP | 60-64 | **< 0.001** | 1.42 | 1.22 – 1.66 | **< 0.001** | 1.30 | 1.14-1.49 |
|  | MAP | 60-64 | **< 0.001** | 1.33 | 1.17 – 1.52 | **< 0.001** | 1.32 | 1.16-1.49 |
|  | SBP | 43 | **0.023** | 1.16 | 1.02 – 1.33 | 0.080 | 1.12 | 0.99-1.26 |
|  | SBP | 53 | **0.004** | 1.22 | 1.06 – 1.39 | **0.006** | 1.19 | 1.05-1.35 |
|  | SBP | 60-64 | **< 0.001** | 1.40 | 1.21 – 1.63 | **< 0.001** | 1.33 | 1.16-1.52 |
| **Cognition** | Cognitive test score | 15 | **0.034** | 0.86 | 0.75 – 0.99 | **0.033** | 0.87 | 0.77-0.99 |
| **Diet** | Canned fruit | 43 | **0.002** | 0.79 | 0.69 – 0.91 | **0.004** | 0.82 | 0.72-0.94 |
|  | Processed pork | 53 | **0.019** | 1.28 | 1.05 – 1.58 | 0.084 | 1.15 | 0.98-1.36 |
|  | Sugar | 53 | **0.029** | 1.24 | 1.02 – 1.50 | 0.226 | 1.10 | 0.94-1.28 |
| **Renal** | Urine creatinine | 60-64 | **0.048** | 0.85 | 0.73 – 1.00 | **0.048** | 0.87 | 0.75-0.99 |
| **Social Determinants** | Dwelling upkeep | 11 | **0.04** | 0.78 | 0.61 – 0.99 | **0.022** | 0.77 | 0.61-0.96 |
|  | Household crowding | 2 | **0.001** | 1.24 | 1.09 – 1.41 | **0.002** | 1.21 | 1.07-1.37 |
|  | Household crowding | 8 | **0.007** | 1.19 | 1.05 – 1.36 | **0.006** | 1.18 | 1.05-1.33 |
|  | Father’s SEP  0 = Manual  1 = Non-manual | 11 | **0.040** | 0.75 | 0.56 – 0.99 | 0.052 | 0.77 | 0.60-1.00 |
|  | Home ownership | 2-11 | **0.007** | 0.78 | 0.65 – 0.93 | **0.011** | 0.81 | 0.68-0.95 |
|  | Housing quality index | 4 | **0.005** | 0.90 | 0.83 – 0.97 | **0.035** | 0.93 | 0.87-0.99 |
|  | Self-employed/ Employed  0 = Not earning  1 = Earning | 60-64 | **0.028** | 0.75 | 0.58 – 0.97 | **0.008** | 0.72 | 0.56-0.92 |

**Table S3**: Significant environmental factors associated with LVDD following multivariable logistic regression adjusting for sex, BMI and SEP at a false discovery rate of 5% in complete case analysis and after multiple imputation. Predictive mean MI was used to generate missing exposures and covariates under the assumption of missingness at random. Categorical variables have their categories listed beneath them.

*Abbreviations: D/SBP = diastolic/systolic blood pressure, GHQ = General Health Questionnaire, MAP = mean arterial pressure, SEP = Socioeconomic position.*

**Table S4**: Significant environmental factors associated with LVDD following multivariable logistic regression adjusting for sex, BMI and SEP in complete case analysis and after participants with a formal cardiovascular disease diagnosis were removed from analysis. Categorical variables have their categories listed beneath them.

|  |  |  | **Complete Case Analysis** | | | **Analysis without CVD** | | |
| --- | --- | --- | --- | --- | --- | --- | --- | --- |
| **Domain** | **Variable** | **Age (years)** | **P-value** | **Odds ratio** | **95% CI** | **P-value** | **Odds ratio** | **95% CI** |
| **Biomarker** | Von Willebrand factor | 60-64 | **0.023** | 1.26 | 1.04 – 1.54 | **0.039** | 1.26 | 1.02-1.57 |
| **Cardiac** | Hypertension | 53 | **0.047** | 1.38 | 1.00 – 1.90 | **0.025** | 1.50 | 1.05-2.12 |
|  | Hypertension | 60-64 | **0.001** | 1.57 | 1.20 – 2.06 | **< 0.001** | 1.67 | 1.24-2.24 |
|  | DBP | 53 | **0.001** | 1.25 | 1.09 – 1.43 | **< 0.001** | 1.31 | 1.13-1.52 |
|  | DBP | 60-64 | **< 0.001** | 1.42 | 1.22 – 1.66 | **< 0.001** | 1.53 | 1.29-1.81 |
|  | MAP | 60-64 | **< 0.001** | 1.33 | 1.17 – 1.52 | **< 0.001** | 1.41 | 1.22-1.65 |
|  | SBP | 43 | **0.023** | 1.16 | 1.02 – 1.33 | **0.019** | 1.19 | 1.03-1.38 |
|  | SBP | 53 | **0.004** | 1.22 | 1.06 – 1.39 | **0.012** | 1.21 | 1.22-1.70 |
|  | SBP | 60-64 | **< 0.001** | 1.40 | 1.21 – 1.63 | **< 0.001** | 1.44 | 1.16-1.52 |
| **Cognition** | Cognitive test score | 15 | **0.034** | 0.86 | 0.75 – 0.99 | **0.021** | 0.84 | 0.72-0.97 |
| **Diet** | Canned fruit | 43 | **0.002** | 0.79 | 0.69 – 0.91 | **0.004** | 0.80 | 0.68-0.93 |
|  | Processed pork | 53 | **0.019** | 1.28 | 1.05 – 1.58 | **0.045** | 1.25 | 1.01-1.57 |
|  | Sugar | 53 | **0.029** | 1.24 | 1.02 – 1.50 | **0.044** | 1.24 | 1.00-1.53 |
| **Renal** | Urine creatinine | 60-64 | **0.048** | 0.85 | 0.73 – 1.00 | 0.194 | 0.89 | 0.75-1.06 |
| **Social Determinants** | Dwelling upkeep | 11 | **0.04** | 0.78 | 0.61 – 0.99 | **0.023** | 0.74 | 0.57-0.96 |
|  | Household crowding | 2 | **0.001** | 1.24 | 1.09 – 1.41 | **0.004** | 1.23 | 1.07-1.42 |
|  | Household crowding | 8 | **0.007** | 1.19 | 1.05 – 1.36 | **0.010** | 1.20 | 1.04-1.38 |
|  | Father’s SEP  0 = Manual  1 = Non-manual | 11 | **0.040** | 0.75 | 0.56 – 0.99 | **0.012** | 0.68 | 0.50-0.92 |
|  | Home ownership | 2-11 | **0.007** | 0.78 | 0.65 – 0.93 | **0.048** | 0.82 | 0.68-1.00 |
|  | Housing quality index | 4 | **0.005** | 0.90 | 0.83 – 0.97 | **< 0.001** | 0.87 | 0.80-0.94 |
|  | Self-employed/ Employed  0 = Not earning  1 = Earning | 60-64 | **0.028** | 0.75 | 0.58 – 0.97 | 0.058 | 0.76 | 0.57-1.01 |

*Abbreviations: CVD = cardiovascular disease, D/SBP = diastolic/systolic blood pressure, GHQ = General Health Questionnaire, MAP = mean arterial pressure, SEP = Socioeconomic position.*

**Figure S1:** Exposome correlation matrices for LVDD spanning childhood (**a**), early adulthood (**b**), middle age (**c**), and older age (**d**). The names of the individual exposures are vertically positioned on the left while the domains are horizontally on top. The strength of the association can be understood using the key on the right.

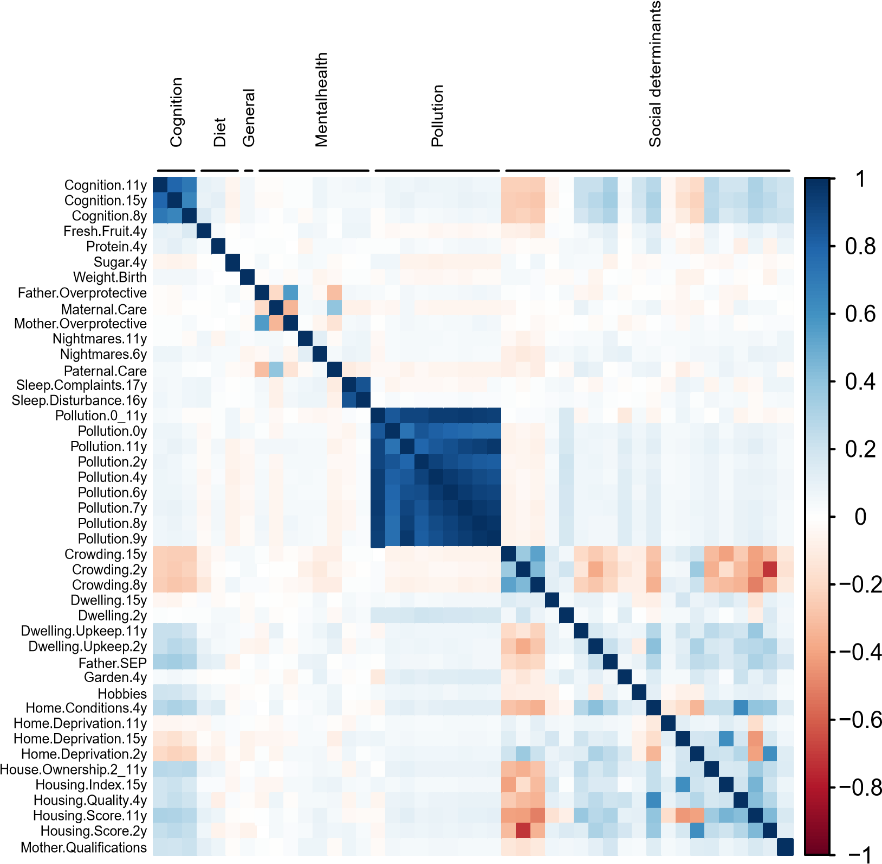

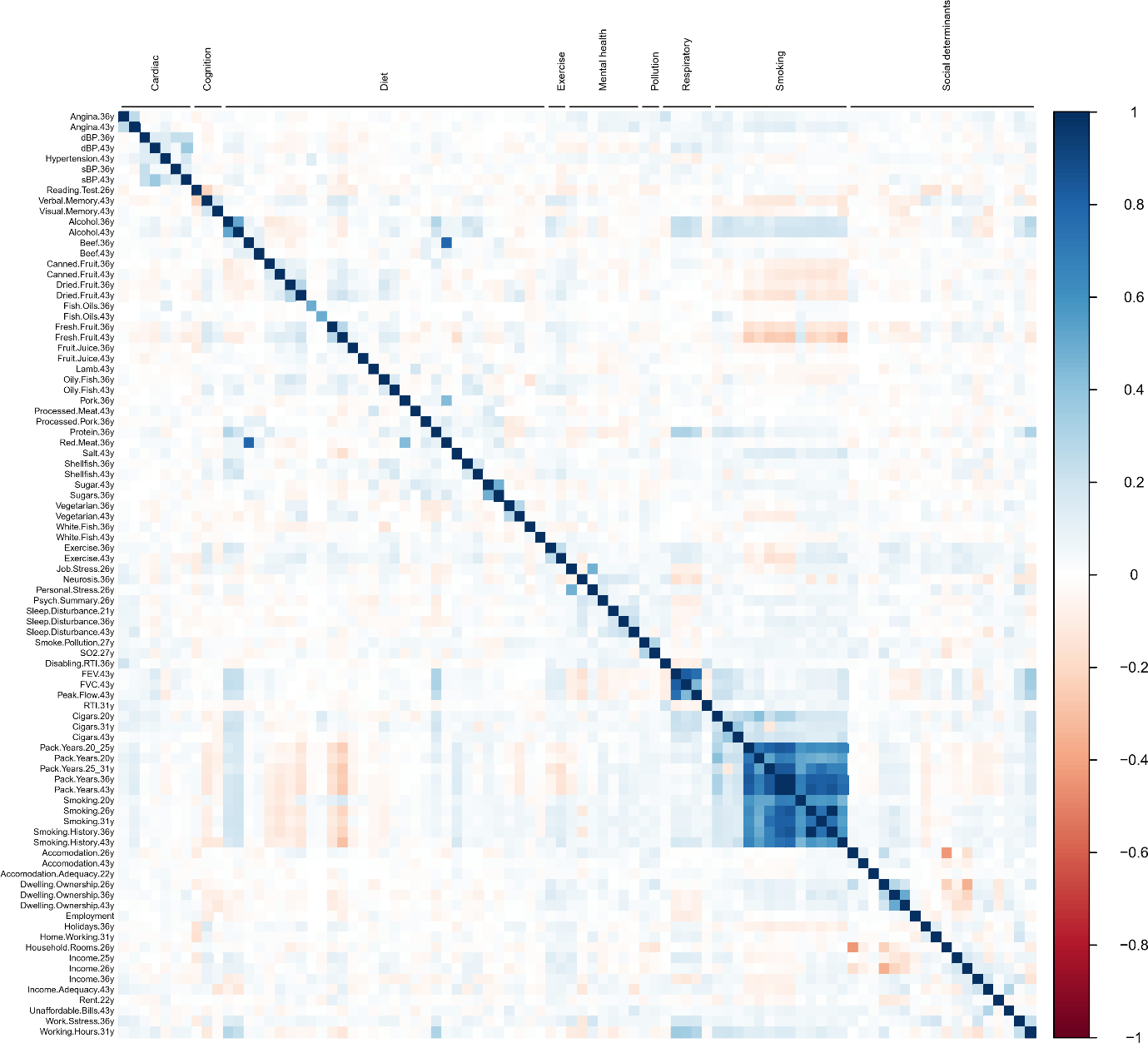

**a)**

**b)**

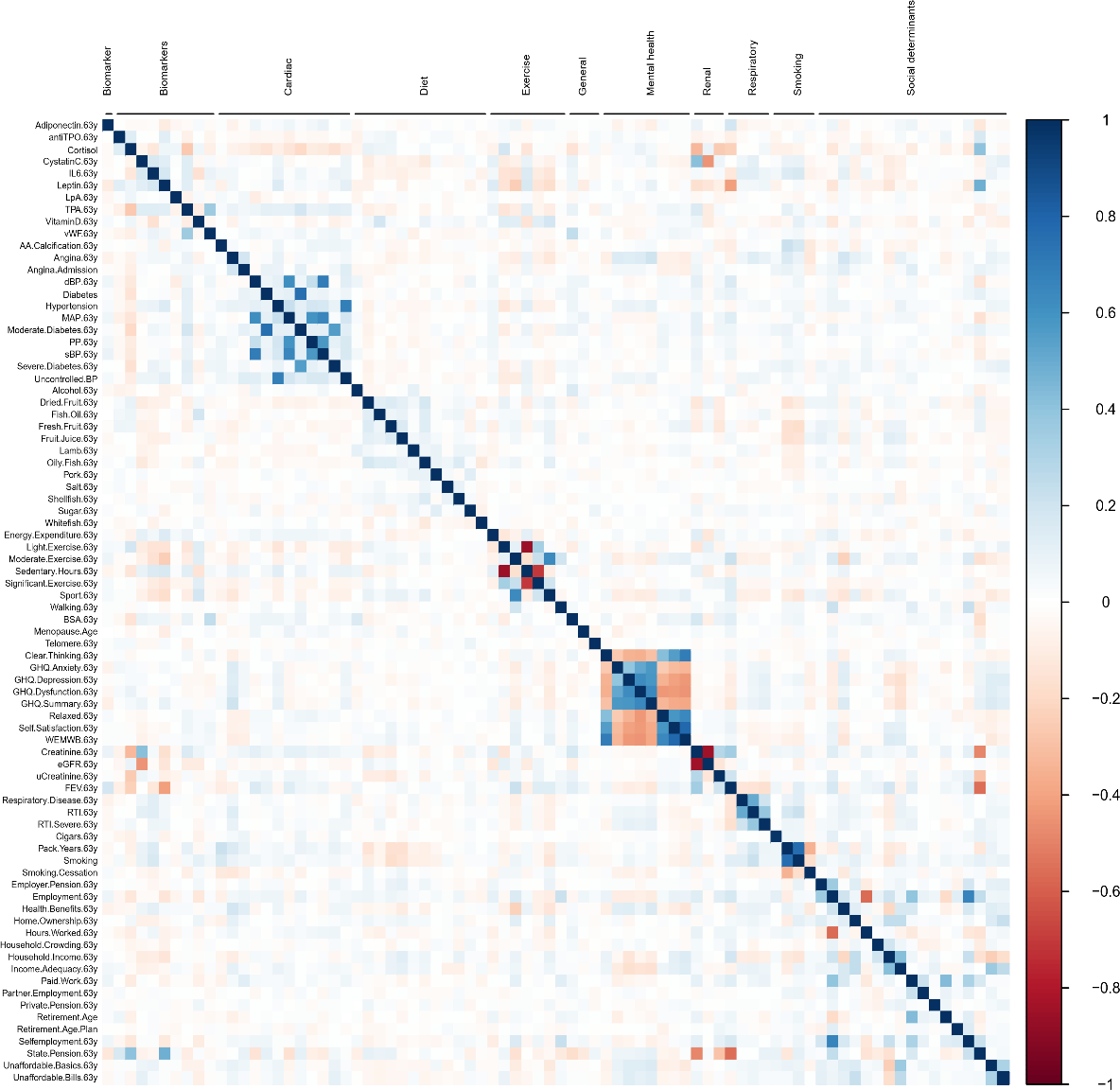

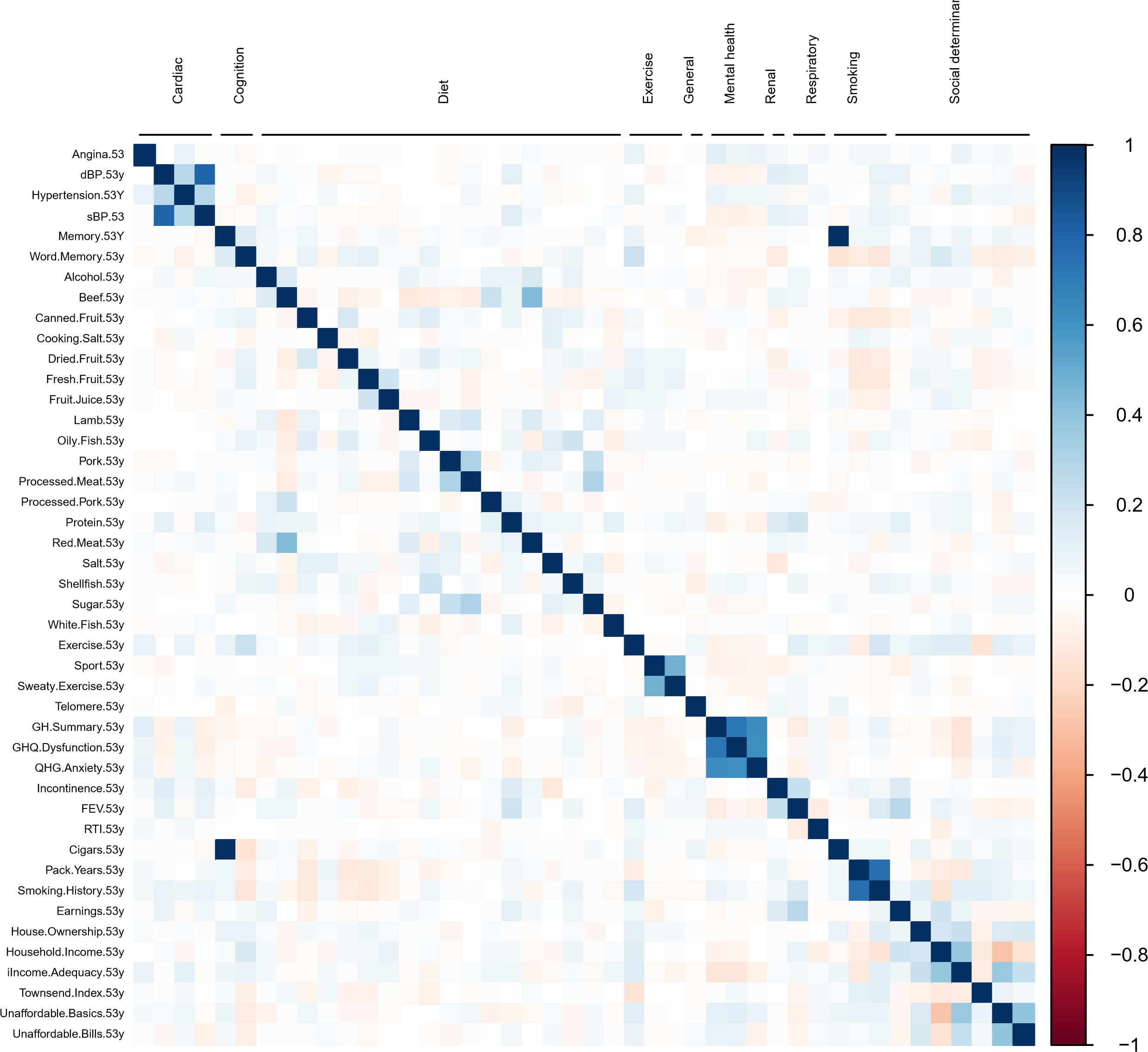

**c)**

**d)**

*Abbreviations: AA.calcification = abdominal aorta calcification, BSA = body surface area, dBP = diastolic blood pressure, FEV = forced expiratory volume, FVC = forced vital capacity, GHQ = general health questionnaire, IL6 = interleukin-6, LpA = lipoprotein A, MAP = mean arterial pressure, PP = pulse pressure, RTI = respiratory tract infection, sBP = systolic blood pressure, SEP = socioeconomic position, TPA = tissue plasminogen activator, TPO = thyroid peroxidase, uCreatinine = urine creatinine, vWF = von Willebrand factor, WEMWB = Warwick-Edinburgh mental wellbeing scale.*
